# In vivo structural MRI-based atlas of human thalamic nuclei

**DOI:** 10.1101/2020.08.09.20171314

**Authors:** Manojkumar Saranathan, Charles Iglehart, Martin Monti, Thomas Tourdias, Brian Rutt

**Author notes:** Corresponding author(s): Manojkumar Saranathan.

## Abstract

Thalamic nuclei play critical roles in regulation of neurological functions like sleep and wakefulness. They are increasingly implicated in neurodegenerative and neurological diseases such as multiple sclerosis and essential tremor. However, segmentation of thalamic nuclei is difficult due to their poor visibility in conventional MRI scans. Sophisticated methods have been proposed which require specialized MRI acquisitions and complex post processing. There are very few digital MRI thalamic atlases and they have been constructed using a small number of post-mortem brains. The goal of this work is the development of a structural thalamic atlas at high spatial resolution based on manual segmentation of 20 subjects that include healthy subjects and patients with multiple-sclerosis. Using data analysis from healthy subjects as well as patients with multiple-sclerosis and essential tremor and at 3T and 7T MRI, we demonstrate the utility of this atlas to provide fast and accurate segmentation of thalamic nuclei when only conventional T1 weighted images are available.

## Background and summary

The thalamus has historically been considered a relay organ, filtering and relaying sensory and motor signals to the cortex. It is also involved in the regulation of sleep, attention, waking, consciousness^1^, and episodic memory^2^. Histologically and functionally, the thalamus is divided into subdivisions called nuclei with specific projections to different cortical areas and associated with specific neurological functions. As a result, thalamus nuclei involvement is increasingly reported in a number of neurodegenerative and psychiatric disorders such as multiple sclerosis^3,4,5^, alcohol use disorder^6^, schizophrenia^7^, and Parkinson’s disease^8^ among others. Specific nuclei such as the ventralis intermedius nucleus are being targeted for treatment of essential tremor^9^. However, thalamic nuclei are largely invisible on conventional T_1_ or T_2_ weighted MRI sequences. Specialized techniques such as susceptibility weighted imaging^10,11^ have been demonstrated at 7T for delineation of thalamic nuclei, although involving tedious manual segmentation and usually focusing primarily on the ventralis intermedius nucleus, a popular target for deep brain stimulation. Diffusion tensor imaging (DTI) has shown promise for delineation of thalamic nuclei. Local^12,13^ properties such as orientation of the diffusion tensor have been utilized to segment the thalamic nuclei into multiple nuclei. To date, the most consistent and stable DTI-based technique uses orientation distribution functions of a spherical harmonic basis to cluster the thalamic nuclei^14^. However, due to the limited spatial resolution and distortion of the underlying echo-planar imaging sequence, and the predominance of grey matter in the thalamus which reduces the anisotropy, DTI-based methods have been successful in only segmenting the larger nuclei.

Very few MRI atlases for thalamic nuclei have been reported. Behrens et al.^15,16^ used probabilistic tractography to create an atlas with seven sub-regions. However, this atlas is based on structural connectivity to the cortex rather than anatomical correspondence to a histological atlas. While the Krauth atlas^17^ is a digital representation of the Morel stereotactic atlas^18^, it is built using 3 healthy *post-mortem* brains. The probabilistic atlas of Iglesias et al^19^ is also, primarily, based on 6 *post-mortem* brains. Recently Najdenovska et al.^20^ reported an atlas based on the DTI clustering method of Battistella et al ^14^ using 70 healthy subjects. This atlas also had seven clusters, six of which loosely corresponded to larger thalamic nuclei while the seventh cluster was a conglomerate of three histologically-defined nuclei. Even though a qualitative correspondence to the Morel atlas was noted, there were no direct quantitative comparisons to manual segmentation ground truth.

Structural MRI is usually performed at much higher spatial resolution than EPI-based methods like DTI and would be ideal for atlas creation. However, T_1_ weighted Magnetization Prepared Rapid Gradient Echo (MP-RAGE) or T_2_weighted fast spin echo structural imaging sequences possess very little inter-nuclear contrast to be of value in nuclei segmentation. Recently, a method for thalamic segmentation called Thalamus optimized multi atlas segmentation (THOMAS)^21^ based on a variant of MP-RAGE has been proposed which shows great promise for high resolution thalamic nuclei segmentation. However, THOMAS requires the acquisition of a white-matter-nulled (WMn) MP-RAGE sequence^22,23^, which has not generally been part of the suite of standard MRI sequences. This also prevents retrospective analysis of large databases like Alzheimer’s Disease Neuroimaging Initiative (ADNI), which have only conventional structural imaging sequences like MP-RAGE.

The goal of this work was to create a high spatial resolution (1 mm^3^) structural atlas based on a database of WMn-MP-RAGE data comprising a mixture of 20 healthy controls and patients with multiple sclerosis, which were segmented manually using the Morel stereotactic atlas as a guide. This allowed delineation of thalamic nuclei from conventional MP-RAGE, enabling their segmentation from existing standard clinical imaging protocols. We describe the creation of this atlas and demonstrate its utility using 3T and 7T MRI data sets.

## Methods

### Datasets and manual segmentation

The structural atlas proposed in this work was generated using 20 WMn-MP-RAGE prior datasets (n=11 subjects (same priors as used in the THOMAS method of Su et al^21^.) with multiple sclerosis, n=9 healthy subjects, mean age= 33.6 years) acquired on a GE 7T MRI system with the following parameters: 180 coronal slices, TR/TE 6,000/10 ms, inversion time 680 ms, flip angle 4°, 1 mm^3^ isotropic resolution, FOV 180 mm, parallel imaging factor 1.5×1.5 (6 datasets with no parallel imaging)

Manual segmentation was performed by an expert neuroradiologist using the Morel atlas^18^ as a reference. A reproducible manual segmentation protocol was developed with excellent intra-rater reliability as measured by intraclass correlation coefficient (ICC) and mean distance discrepancy between centers of mass (ΔCoMs) for initial and repeat tracings 3 weeks later, yielding ICC of 0.997 (95% confidence interval 0.996–0.998) and ΔCoM of 0.69 ± 0.38 mm respectively. More details of the manual segmentation procedure can be found in Tourdias et al.^22^. All 20 prior datasets were manually segmented to identify 11 thalamic nuclei and the mammillothalamic tract (MTT). The eleven delineated nuclei are grouped as follows:

i. **anterior group:** anteroventral (AV)
ii. **lateral group:** ventral posterolateral (VPL), ventral lateral anterior (VLa), ventral lateral posterior (VLp), ventral anterior nucleus (VA)
iii. **medial group:** mediodorsal (MD), centromedian (CM), habenula (Hb)
iv. **posterior group:** pulvinar (Pul), medial geniculate nucleus (MGN), lateral geniculate nucleus (LGN)

### Custom template construction and atlas creation

A custom template was created using the buildtemplate script of Advanced Normalization Tools (ANTs^24^) as described in Su et al^21^. Briefly, this is achieved by iteratively registering each of the 20 priors to the average of the priors and then averaging the registered priors to create a custom template which has very high signal-to-noise ratio and contrast whilst including normal and diseased brain states. Registration was affine followed by nonlinear, with the symmetric group-wise diffeomorphic normalization (SyN) algorithm of ANTs was used for all nonlinear warping. ANTs was chosen for its accuracy and precision as reported by Klein et al ^25^. The nonlinear warps from each prior to the custom template were also computed using ANTs. Finally, labels were transferred from the space of the 20 priors to the custom template space using the warps computed above and nearest-neighbor interpolation to generate the thalamic parcellations in template space. These 20 parcellations were then combined to calculate the spatial probability maps and maximum probability map using custom python scripts. Spatial probability maps were generated by computing the relative frequencies of labels at each voxel in template space to yield the probability of that voxel belonging to each thalamic nucleus. Maximum probability maps were computed using the mode of these distributions, thus assigning a single label to each voxel representative of the most probable thalamic nucleus at that location. Lastly, the custom template was nonlinearly registered to MNI space (nonlinear ICBM152 asymmetric) and this spatial warp was saved and used to warp the probability maps from custom template to MNI space. These steps are summarized in Figure 1.

**Figure 1.**
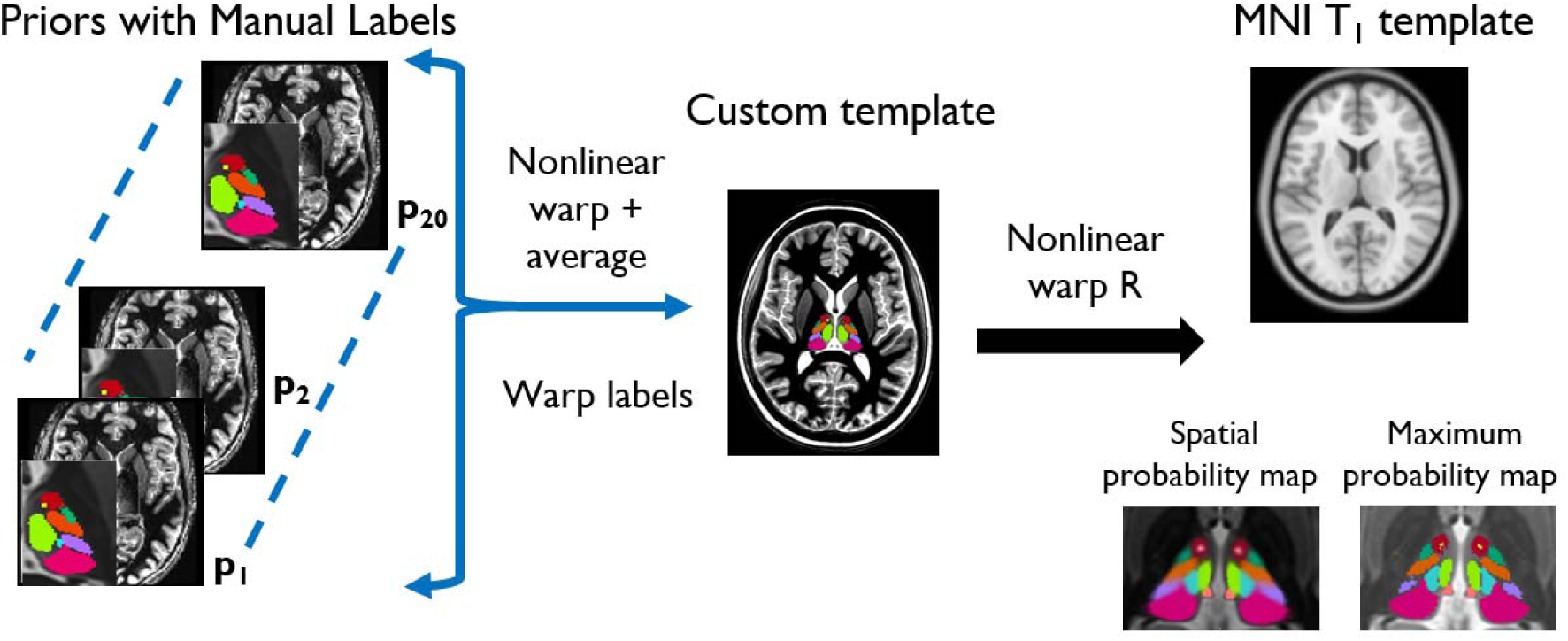
Main steps in the creation of the proposed thalamic atlas

## Data Records

The primary contribution of our work are the spatial probability and maximum probability maps of thalamic nuclei in custom template space. They are in compressed NIfTI-1 format (a *.nii.gz* extension) with the spatial probability map a 4-D file, the 4^th^ dimension of size 24 for the 12 left and 12 right thalamic nuclei and separate maximum probability maps for left and right thalami. The maximum probability maps are also provided in MNI 152 (nonlinear 2009c) space at 0.5 mm^3^ isotropic resolution. Figure 2 shows the spatial probability maps and maximum probability maps overlaid on the custom template in all three planes. The maximum probability map in MNI space is shown in Figure 3. A customized color lookup table (a *.ctbl* extension) recognized by standard visualization tools like 3D Slicer is also provided. This can be custom edited to change the color scheme or add additional nuclei. The data is available through zenodo (Data Citation 1). In addition, code for segmentation of conventional MP-RAGE using the atlas and some extra files used by the code are also provided (see Code availability section). The 20 original WMn-MP-RAGE datasets used for atlas creation and their segmentation as well as their warps to custom template and the custom template are available through github (Data Citation 2) in compressed NlfTI-1 format (i.e. *.nii.gz* extension). A summary of data records is shown in Table 1.

**Table 1.** Summary of data records related to this study

| Dataset | Data for atlas creation | Data used for testing |
| --- | --- | --- |
| Number of subjects | 20 | 36 |
| Provenance | Prior data from Su et al.<sup>19</sup> | Mixed (see Methods) |
| Available from | Github Data Citation 2 | Subset in Data Citation 1 |
| Modalities used | WMn MP-RAGE | WMn MP-RAGE and MP- RAGE |
| Use | Atlas creation | Atlas validation |
| Provided output | Spatial and max. probability maps | THOMAS and atlas-based thalamic nuclei |
| Output format | NifTI-1 (.gz) 4D and 3D | NifTI-1 (.gz) 3D |

**Figure 2.**
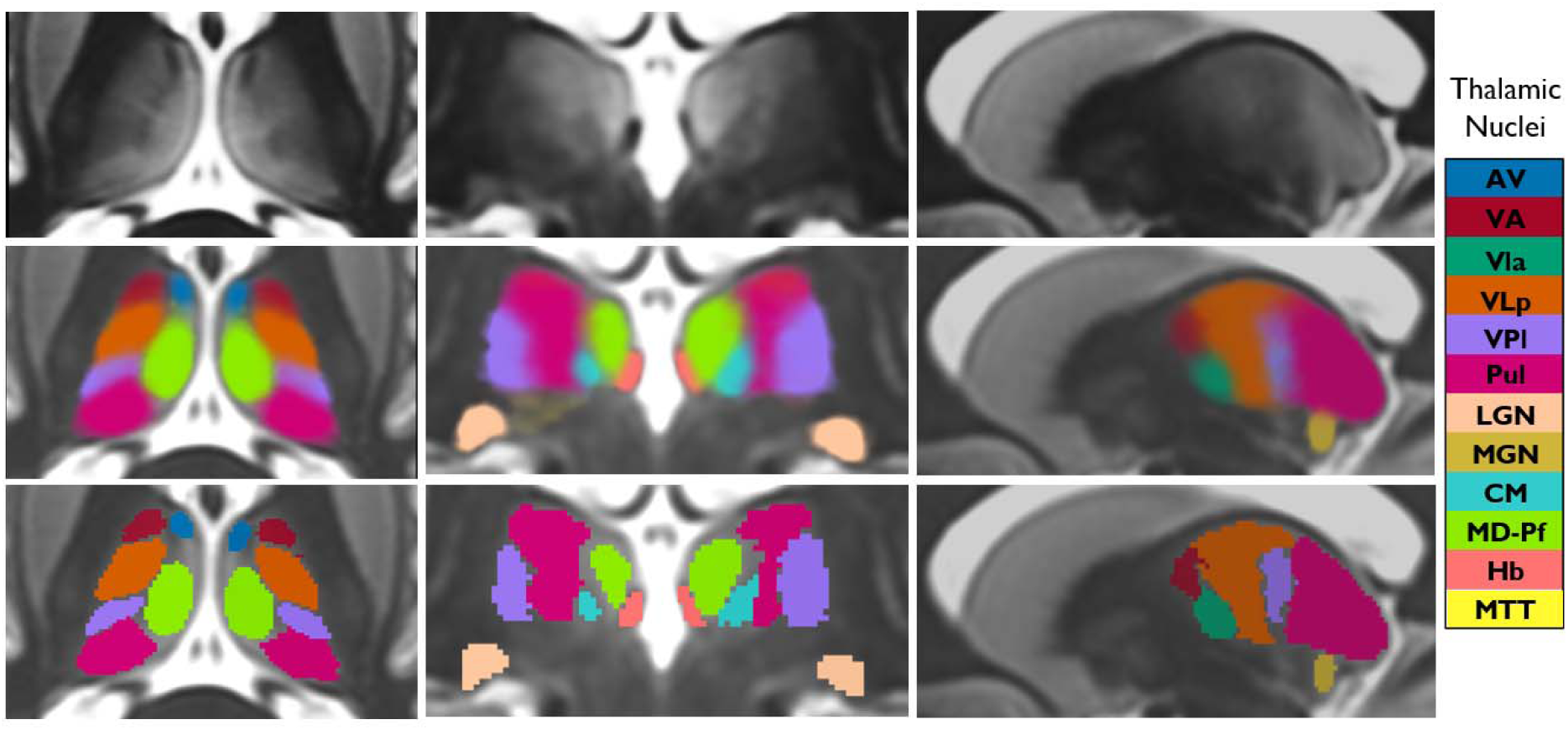
Spatial probability maps (middle row) and maximum probability maps (bottom row) in custom template space shown in three orthogonal planes. The top row shows the custom template without overlays for reference.

**Figure 3.**
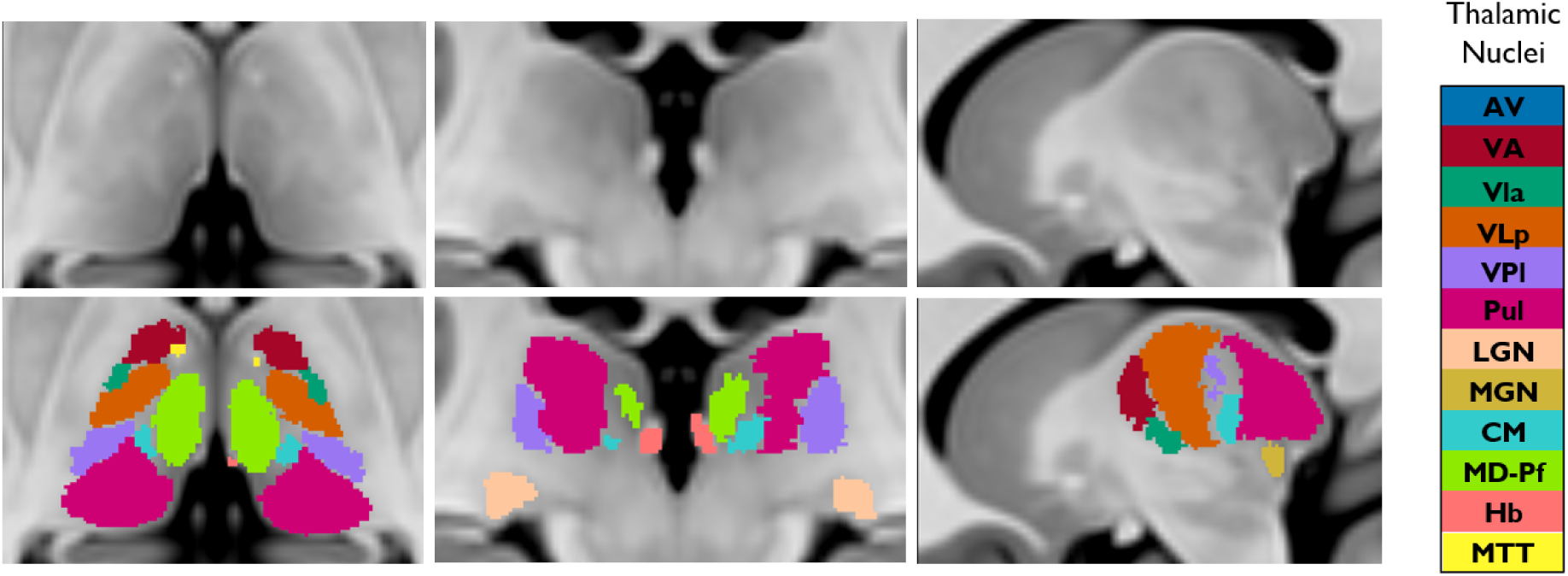
Maximum probability thalamic maps in MNI152 space in three orthogonal planes.

## Technical Validation

To test the accuracy of the proposed atlas-based segmentation method, two datasets were used. The first comprised of data from patients with essential tremor, multiple sclerosis, as well as healthy subjects (total n=18) acquired on a 7T GE scanner using a 32-channel array (Nova Medical Systems). These were completely separate from the 20 prior subjects used for atlas construction. The second test dataset comprised of 18 healthy subjects acquired on a 3T Siemens Prisma scanner using a 32-channel array. All subjects were scanned after written informed consent adhering to institutional review board (IRB) guidelines. The scan parameters for the sequences were as follows:

**7T:** *Conventional MP-RAGE-*180 coronal slices, TR/TE 3,000/7.2 ms, flip angle 6°, inversion time 1200 ms, 1mm isovoxel resolution, Field of view (FOV) 180 mm, Autocalibrating reconstruction for Cartesian imaging (ARC) acceleration factor 2

*WMn MP-RAGE-*180 coronal slices, TR/TE 6,000/10 ms, inversion time 680 ms, flip angle 4°, 1mm isotropic resolution, FOV 180 mm, ARC factor 1.5×1.5

**3T:** *Conventional MP-RAGE-*192 sagittal slices, TR/TE 2,000/2.52 ms, flip angle 12°, 1mm isovoxel resolution, FOV 256 mm, generalized autocalibrating partially parallel acquisitions (GRAPPA) factor 2 *WMn MP-RAGE:* 160 axial slices, TR/TE 4,000/3.75 ms, inversion time 500 ms, flip angle 7°, 1mm isotropic resolution, FOV 256 mm, GRAPPA factor 2

For both 7T and 3T datasets, WMn MP-RAGE images from each patient were segmented using THOMAS and conventional MP-RAGE using the proposed atlas-based segmentation approach, respectively. For the 7T data, manual segmentation on WMn MP-RAGE performed by a trained neuroradiologist guided by the Morel atlas were also available. As a result, THOMAS and the atlas-based segmentations were individually compared to the manual segmentation ground truth. For the 3T data set, the atlas-based segmentation was directly compared to THOMAS segmentation, due to lack of manual segmentation ground truth. The WMn and conventional MP-RAGE data from each patient were affine registered prior to quantitative comparisons. Their associated labels were also registered by applying the same affine transform with nearest neighbor interpolation. Figure 4 shows comparison of THOMAS segmentation on WMn-MP-RAGE (left column) with atlas-based segmentation on conventional MP-RAGE (right column) for a MS patient at 7T (top row) and a healthy subject at 3T (bottom row). The qualitative agreement of the methods can be appreciated.

**Figure 4.**
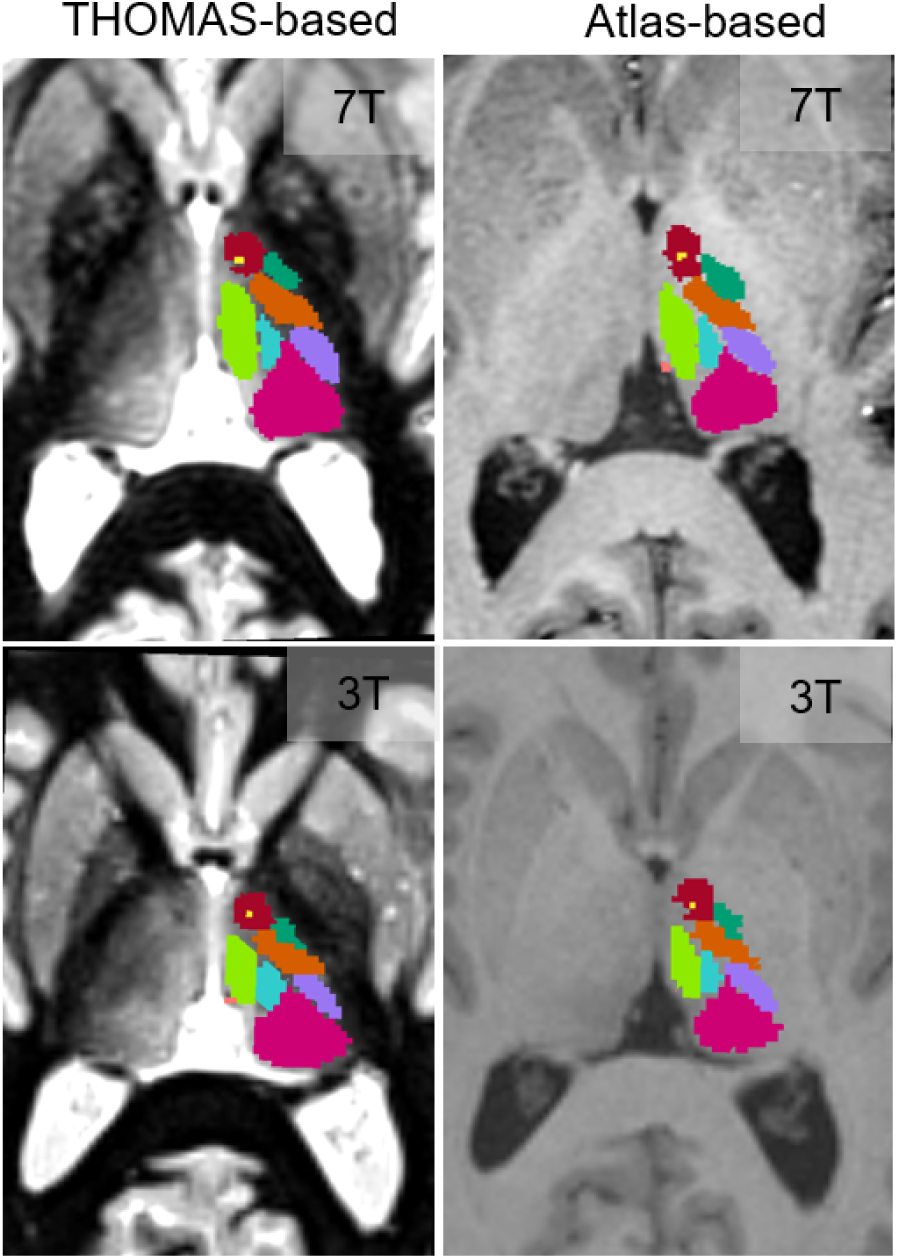
Comparison of THOMAS-based and the proposed atlas-based segmentation on a MS patient at 7T (top row) and a healthy subject at 3T (bottom row)

The main quantitative measures used for comparisons were Dice coefficients and Volume Similarity Index (VSI). These are defined as

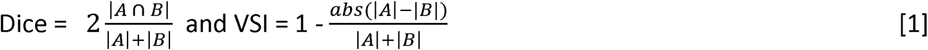

where A and B refer to the two segmentation labels compared and |A| and |B| refers to the number of pixels in A and B respectively

For 3T, the difference between the centroids of the atlas and THOMAS labels was also computed for each of the 11 segmented nuclei.

### Results

Dice and VSI for the 7T test data are shown in Table 1 for the whole thalamus and 11 segmented nuclei. For THOMAS segmentation, mean Dice and VSI were 0.74 and 0.92 for the larger nuclei (shaded rows) and 0.64 and 0.83 for the smaller nuclei (unshaded rows) using THOMAS. For the atlas-based segmentation, mean Dice and VSI were 0.68 and 0.92 for the larger nuclei (shaded rows) and 0.58 and 0.88 for the smaller nuclei (unshaded rows). While there is a slight reduction in Dice for the atlas-based method, especially for the smaller nuclei, the reductions are under 10% except for CM and Hb (~14%). VSI was comparable for most nuclei.

Dice and VSI for the 3T test data are shown in Table 2 for the whole thalamus and 11 segmented nuclei. Note that these are in comparison to THOMAS as opposed to a manual gold standard. The mean Dice and VSI were 0.8 and 0.95 for the larger nuclei (shaded rows) and 0.7 and 0.91 for the smaller nuclei (unshaded rows), indicating a fairly high degree of concordance. The distance between the centroids was less than 1 mm for all nuclei except AV, averaging 0.67 mm and 0.75 mm for the larger and smaller nuclei respectively, further attesting to the accuracy of the atlas-based method compared to THOMAS.

**Table 2.** Dice and VSI values for 7T test data. * indicates p<0.05

| Nucleus | Dice | Dice | VSI | VSI |
| --- | --- | --- | --- | --- |
|  | THOMAS vs. manual | Atlas vs. manual | THOMAS vs. manual | Atlas vs. manual |
| *Whole thalamus* | 0.89 ± 0.02* | 0.88 ± 0.02 | 0.95 ± 0.04 | 0.95 ± 0.04 |
| *Pulvinar (Pul)* | 0.84 ± 0.03* | 0.79 ± 0.04 | 0.95 ± 0.02 | 0.94 ± 0.05 |
| *Ventrolateral posterior (VLp)* | 0.77 ± 0.04* | 0.70 ± 0.05 | 0.92 ± 0.06 | 0.92 ± 0.06 |
| *Mediodorsal (MD)* | 0.83 ± 0.04* | 0.77 ± 0.06 | 0.92 ± 0.05 | 0.90 ± 0.06 |
| *Ventral posterior lateral (VPI)* | 0.63 ± 0.12 | 0.59 ± 0.11 | 0.89 ± 0.08 | 0.94 ± 0.05 |
| *Ventral Anterior (VA)* | 0.62 ± 0.09* | 0.57 ± 0.10 | 0.93 ± 0.04 | 0.92 ± 0.06 |
| *Anteroventral (AV)* | 0.66 ± 0.15 | 0.59 ± 0.06 | 0.75 ± 0.16 | 0.86 ± 0.09 |
| *Centromedian (CM)* | 0.68 ± 0.14* | 0.59 ± 0.17 | 0.92 ± 0.07* | 0.89 ± 0.08 |
| *Lateral geniculate nucleus (LGN)* | 0.58 ± 0.17 | 0.53 ± 0.14 | 0.85 ± 0.12 | 0.91 ± 0.07 |
| *Ventral lateral anterior (VLa)* | 0.52 ± 0.12 | 0.51 ± 0.15 | 0.75 ± 0.16 | 0.89 ± 0.07* |
| *Medial geniculate nucleus (MGN)* | 0.64 ± 0.09 | 0.63 ± 0.08 | 0.80 ± 0.13 | 0.85 ± 0.10* |
| *Habenula (Hb)* | 0.76 ± 0.05* | 0.65 ± 0.08 | 0.90 ± 0.07 | 0.87 ± 0.09 |

**Table 3.** Dice and VSI values for 3T test data.

| Nucleus | Dice<br>Atlas vs.<br>THOMAS | VSI<br>Atlas vs.<br>THOMAS | Distance<br>between<br>centroids<br>(mm) |
| --- | --- | --- | --- |
| *Whole thalamus* | 0.92 ± 0.01 | 0.98 ± 0.01 | 0.54 |
| *Pulvinar (Pul)* | 0.86 ± 0.02 | 0.96 ± 0.03 | 0.60 |
| *Ventrolateral posterior (VLp)* | 0.82 ± 0.03 | 0.97 ± 0.02 | 0.67 |
| *Mediodorsal (MD)* | 0.86 ± 0.03 | 0.97 ± 0.02 | 0.66 |
| *Ventral posterior lateral (VPI)* | 0.70 ± 0.08 | 0.89 ± 0.05 | 0.83 |
| *Ventral Anterior (VA)* | 0.77 ± 0.03 | 0.96 ± 0.03 | 0.61 |
| *Anteroventral (AV)* | 0.67 ± 0.07 | 0.89 ± 0.07 | 1.26 |
| *Centromedian (CM)* | 0.72 ± 0.05 | 0.94 ± 0.02 | 0.75 |
| *Lateral geniculate nucleus (LGN)* | 0.75 ± 0.06 | 0.92 ± 0.05 | 0.64 |
| *Ventral lateral anterior (VLa)* | 0.66 ± 0.07 | 0.80 ± 0.07 | 0.75 |
| *Medial geniculate nucleus (MGN)* | 0.76 ± 0.05 | 0.94 ± 0.03 | 0.49 |
| *Habenula (Hb)* | 0.67 ± 0.08 | 0.95 ± 0.04 | 0.62 |

## Usage notes

The atlases provided are in slice correspondence with the standard MNI 152 nonlinear 2009c atlases. Code is also provided for users to efficiently derive thalamic parcellation of their input data using the supplied templates and atlases. A readme file explains the different files and their roles.

## Code availability

The code for the segmentation is a shell script which is provided in the zenodo distribution (Data citation 1). It performs an automatic cropping of the input dataset prior to registering to a cropped custom template. This is done to speed up registration and for accuracy by focusing on the thalami as the crop region encompasses both thalami. A mask for automatic cropping and the cropped custom template are also provided.

## Data Availability

The outputs i.e. probabilistic atlases are available on zenodo along with code and also a small part of the test data. https://zenodo.org/record/3966531

## Author contributions

MS-co-designed the study, programmed the segmentation method, analyzed the data, wrote the manuscript

CI-helped with data analysis and figure preparation

TT-interpretation and discussions, revised the manuscript

MM-interpretation and discussions, revised the manuscript

BR-co-designed the study, revised the manuscript

## Competing interests

No competing interests to report for any of the authors.

## Notes

### Competing Interest Statement

The authors have declared no competing interest.

### Funding Statement

No external funding was received

### Author Declarations

Stanford and UCLA IRB

